# A Shift Toward Proteolytic Gut Fermentation Links Systemic Inflammation to Clinical Phenotypes in Major Depressive Disorder

**DOI:** 10.64898/2026.03.22.26348995

**Authors:** Mengqi Niu, Yiping Luo, Chenkai Yangyang, Abbas F. Almulla, Andre F. Carvalho, Jing Li, Yingqian Zhang, Michael Maes

## Abstract

**Background:** The "Neuro-Immune-Metabolic-Oxidative Stress" (NIMETOX) theory identified systemic dysregulation in Major Depressive Disorder (MDD), yet the precise gut-derived metabolic triggers initiating this cascade remain elusive. This study investigated the interplay between fecal short-chain fatty acids (SCFAs), systemic immune activation, and clinical phenotypes to identify a potential "gut-immune biotype" for MDD.

**Methods:** Fecal SCFA profiles and serum immune-inflammatory markers were quantified in 102 patients with MDD and 38 matched healthy controls. A multistage statistical approach was employed: binary logistic regression and 10-fold cross-validated linear discriminant analysis (LDA) were utilized to evaluate diagnostic accuracy, while multivariable regression models were applied to identify robust predictors of clinical phenotypes, including the overall severity of depression (OSOD), physiosomatic symptoms, and recurrence of illness (ROI).

**Results:** MDD patients exhibited a significant depletion of protective straight-chain SCFAs (acetate, propionate, butyrate) and an elevation in branched-chain SCFAs (BSCFAs), indicating a pathological shift from saccharolytic to proteolytic fermentation. This metabolic shift correlated with elevated acute phase-inflammatory index (API) and epidermal growth factor (EGF). A multidimensional model combining BSCFAs, acetate, API, EGF, and T helper 2 discriminated MDD from controls with adequate accuracy (AUC = 0.874). Furthermore, elevated BSCFAs and decreased protective SCFAs strongly predicted higher OSOD, more severe physiosomatic symptoms, and increased ROI. Notably, 5-Hydroxytryptamine receptor 1A agonists were independently associated with elevated BSCFAs.

**Conclusion:** MDD is characterized by a distinct "gut-immune biotype" tightly linked to toxic proteolytic gut fermentation. This metabolic-immune fingerprint offers an objective diagnostic tool and highlights the need for microbiome-targeted interventions in precision psychiatry.

## Introduction

Major Depressive Disorder (MDD) is a leading cause of global disability, affecting approximately 5.7% of adults and imposing a substantial societal burden (1). Due to its elusive pathophysiology, diagnosis relies on subjective symptoms rather than objective biomarkers, resulting in high heterogeneity, delayed treatment, and variable outcomes (2). Recent reviews have proposed the concept of the "Neuro-Immune-Metabolic-Oxidative Stress (NIMETOX)” theory of MDD. Transcending the traditional monoamine hypothesis, this theory posits that MDD operates as a vicious cycle characterized by oxidative stress, loss of metabolic flexibility, and a breakdown of immune tolerance (3). However, the upstream drivers initiating this systemic cascade, as well as objective biomarkers reflecting their specific trajectory, remain elusive.

Emerging evidence suggests that the microbiota-gut-brain (MGB) axis is one of the key systems regulating the NIMETOX pathway via effects of leaky gut or increased gut permeability or the production of immune-modifying metabolites (3-8). Often described as a "forgotten endocrine organ," the gut microbiota finely tunes host physiology through its metabolites (4, 9, 10). Among these, short-chain fatty acids (SCFAs)—primarily acetate, propionate, and butyrate produced via the saccharolytic fermentation of dietary fiber—are cornerstones of gut homeostasis and barrier function (11-13). Previous studies have consistently observed a reduction in butyrate-producing bacteria, such as *Faecalibacterium* and *Coprococcus*, in MDD (4, 14, 15). In a healthy state, acetate and butyrate are efficiently absorbed into colonocytes via monocarboxylate transporters (MCT1/SMCT1); specifically, butyrate acts as a primary energy source and reinforces tight junction protein expression by activating GPR109A and FFAR2 receptors (16, 17). By fortifying these barriers, SCFAs restrict the translocation of inflammatory molecules like lipopolysaccharides (LPS), thereby potentially attenuating systemic inflammation and subsequent neuroinflammation (6, 7, 18-20).

However, the current literature has predominantly focused on these beneficial saccharolytic products, largely overlooking other minor yet bioactive fermentation metabolites. This includes not only branched-chain SCFAs (BSCFAs; e.g., isovaleric, isobutyric, and 2-methylbutyric acid) derived from proteolytic fermentation, but also longer-chain carboxylic acids such as pentanoic and hexanoic acids. BSCFAs are primarily produced by proteolytic bacteria such as *Bacteroides*, *Clostridium*, and *Alistipes*, genera frequently reported to be abnormally abundant in patients with MDD (4, 14, 15, 21). Unlike saccharolytic fermentation, the generation of BSCFAs is often accompanied by the accumulation of potent cytotoxic metabolites, including ammonia, phenols (e.g., p-cresol), and hydrogen sulfide (22). This shift signifies not merely a change in metabolic substrates but a pathological transition of the gut microenvironment toward a state of "metabolic toxicity" or dysregulation, which may directly damage the intestinal epithelium and drive systemic inflammation (23, 24). Furthermore, the physiological roles of the trace metabolites pentanoic and hexanoic acids remain a subject of debate. While elevated concentrations of these acids are may be regarded as indicators of gut dysbiosis, emerging evidence suggests that they may have alternative functions. Specifically, valeric acid may act as a physiological epigenetic modulator, offering potential neuroprotective benefits by inhibiting histone deacetylases (HDACs) (25).

Parallel to gut-brain axis dysfunctions, immune-inflammatory dysregulation is a core driver of the NIMETOX pathways (3, 26-31). Patients with depression often exhibit a state of low-grade "sterile inflammation," characterized by sustained elevations in positive acute phase proteins (APPs) (monomeric C-reactive protein), reduced levels of negative APPs (albumin and transferrin), and growth factors such as EGF (26-30). Accumulating evidence suggests this inflammatory state might partly stem from the breakdown of gut barrier function. The depletion of butyrate downregulates the expression of tight junction proteins (e.g., Claudin-5, Occludin), triggering "leaky gut" and facilitating the translocation of bacterial LPS into the circulation (18, 19). Studies have identified elevated IgA and IgM responses against LPS in MDD patients, accompanied by increased permeability markers like Zonulin (6, 7). This translocated LPS can activate the Toll-like receptor 4 (TLR4)/NF-κB pathway, initiating a "TLR-radical cycle" and subsequent chronic activation of the immune-inflammatory response system (IRS) (20). Specifically, translocated LPS increases oxidative stress, generating new damage-associated molecular patterns (DAMPs) that further activate TLRs, creating an auto-amplifying inflammatory loop (20). Furthermore, complementing this pathogen-driven inflammation, the accumulation of specific intermediate metabolites, such as succinic acid (succinate), serves as a crucial immunometabolic signal. Succinate can stabilize HIF-1α in macrophages, driving their polarization toward a proinflammatory M1 phenotype, thereby converging with LPS signaling to exacerbate the inflammatory milieu (32). Nevertheless, how specific imbalances in the fecal metabolic profile—encompassing protective SCFAs, toxic proteolytic products, and immunometabolic intermediates like succinate—specifically perturb immune tolerance remains to be elucidated.

Moreover, activation of the NIMETOX pathway does not occur in isolation but is profoundly influenced by environmental and therapeutic factors (3). Adverse Childhood Experiences (ACEs) are major risk factors for MDD, potentially causing persistent immune sensitization and metabolic abnormalities through "biological imprinting" (33-40). Simultaneously, while the impact of psychotropic medications on the gut microbiome is gaining attention, their specific effects on the full spectrum of SCFA profiles remain underexplored, constituting a significant confounding factor (41-43).

Niu et al. (26) have demonstrated that the phenotype of MDD comprises interconnected domains of mood, somatic, autonomic, melancholic, and suicidal symptoms. The overall severity of depression (OSOD) signifies a general element that underlies various domains, whereas a singular group factor denotes a physiosomatic dimension containing specific fatigue-related and physical symptoms (26). Moreover, the recurrence of illness (ROI) index should be employed to evaluate the clinical characteristics of MDD; ROI measures the lifetime disease burden linked to depressive episodes and suicidal behaviors (44, 45). Currently, there is a lack of comprehensive research integrating fecal SCFAs, immune inflammation, ACEs, and medication effects to evaluate their diagnostic value for MDD and their explanatory power for clinical phenotypes such as OSOD, physiosomatic symptoms, current suicidal ideation (SI), and ROI.

Addressing these gaps, this study aims to: (1) comprehensively analyze the profiles of SCFAs, including straight-chain SCFAs (acetate, propionate, butyrate), BSCFAs (isovaleric, isobutyric, and 2-methylbutyric acid), straight-chain C5–C6 fatty acids (pentanoate, hexanoate), and the intermediate metabolite succinic acid in the feces of MDD patients; (2) evaluate the diagnostic accuracy of a model combining SCFA and immune markers in distinguishing MDD from healthy controls (HCs); and (3) explore how interactions among SCFAs, immune markers, and ACEs predict the key clinical phenotypes. We hypothesize that MDD is characterized by a specific "metabolic-immune fingerprint" (low protective SCFAs, high BSCFAs/dysregulated acids, and high inflammation) and that this fingerprint reflects NIMETOX pathway activation, driving clinical severity.

## Methods

### Participants

This cross-sectional study enrolled 140 participants, including 102 patients with MDD and 38 HCs. Patients were recruited from the Mental Health Center of Sichuan Provincial People’s Hospital (Chengdu, China). All participants were aged 18-65 years. MDD was diagnosed based on DSM-5 criteria and necessitated a 21-item Hamilton Depression Rating Scale (HAMD-21) (46) score exceeding 18. Controls were selected among hospital personnel and their relatives/acquaintances and were matched to patients by sex, age, years of education, and body mass index (BMI).

Participants were excluded from eligibility if they had any neurological condition, including Alzheimer’s disease, Parkinson’s disease, stroke, epilepsy, brain tumors, or multiple sclerosis, or if they had developmental or personality disorders. Subjects were also eliminated if they had significant medical or immune-related illness, such as type 1 diabetes, rheumatoid arthritis, autoimmune disorders, systemic lupus erythematosus, inflammatory bowel disease, chronic obstructive pulmonary disease, psoriasis, or cancer. Furthermore, participants were excluded if they had other major psychiatric disorders, including bipolar disorder, schizoaffective disorder, schizophrenia, organic mental disorders, autism spectrum disorder, or substance use disorders, excluding nicotine dependency. The additional exclusion criteria included pregnancy or breastfeeding, severe allergic reactions in the prior month, infection or surgery within the past 3 months, current treatment with corticosteroids, immunosuppressive agents, or other immunomodulatory medications, use of therapeutic-dose antioxidants or omega-3 supplements during the preceding 3 months, and frequent consumption of analgesics. For healthy controls, further exclusion criteria encompassed any present or prior history of MDD, dysthymia, or DSM-5 anxiety disorders, along with a familial history of affective disorders, suicide, or substance use disorders, excluding nicotine dependency. All participants or their legal guardians provided written informed consent. The Ethics Committee of Sichuan Provincial People’s Hospital approved the study (Ethics (Research) 2024-203).

### Clinical evaluation

A certified physician conducted semi-structured interviews to collect sociodemographic information and medical, psychiatric, and familial histories. Psychiatric diagnoses were verified using the Mini-International Neuropsychiatric Interview (M.I.N.I.) (47), which assesses a broad range of psychiatric conditions. These included anxiety disorders (social anxiety disorder, panic disorder, generalized anxiety disorder, and agoraphobia), mood-related conditions (depressive episodes, hypomanic episodes, and dysthymia), trauma- and obsessive-compulsive-related disorders, psychotic disorders, eating disorders, alcohol- and non-alcohol-related substance abuse/dependence, and antisocial personality disorder.

Symptom severity ratings were completed on the same day by the same assessor. Scales and computation procedures for OSOD, physiosomatic symptoms, current SI, and ROI followed Niu et al (26). ROI was operationalized as a composite z-score obtained by summing lifetime z-scores for the number of depressive episodes, suicide attempts (SA), and suicidal ideation (SI). ACEs included emotional abuse, physical abuse, sexual abuse, emotional neglect, and physical neglect; the sum of the four non-sexual ACE domains was defined as “Four ACEs” (48).

Metabolic syndrome (MetS) components included height, weight, BMI, and waist circumference (WC). BMI was computed as weight (kg) divided by height (m) squared, while WC was assessed at the midpoint between the iliac crest and the lower rib edge. MetS was defined according to the 2009 joint interim statement (AHA/NHLBI) (49): MetS was diagnosed when ≥3 of the following criteria were met: (a) WC ≥90 cm (men) or ≥80 cm (women); (b) triglycerides ≥150 mg/dL; (c) HDL-C <40 mg/dL (men) or <50 mg/dL (women); (d) systolic blood pressure ≥130 mmHg or diastolic blood pressure ≥85 mmHg, or antihypertensive treatment; and (e) fasting glucose ≥100 mg/dL or diagnosed diabetes. We also used a MetS ranking based on the number of criteria met.

### Assays

Between 6:30 and 8:00 a.m., 30 mL of fasting venous blood was collected into serum tubes. Samples were centrifuged at 3500 rpm, and serum was aliquoted and stored at -80°C until analysis. **ESF, Table 1** lists all quantified analytes along with the corresponding methods and instrumentation; **ESF, Table 2** provides the composition and calculation of composite biomarker indices (acute phase-inflammatory index (API), Th1, Th17, Th2, IRS, compensatory immune-regulatory system (CIRS), protective SCFAs, neutral SCFAs, and BSCFA index).

### Data analysis

All analyses were performed using IBM SPSS Statistics 30 for Windows. Data distributions were inspected prior to analysis, and variables were transformed as needed (log10, square root, rank-based, or Winsorized transformations). Group differences in continuous variables were tested using ANOVA, and categorical variables were examined using contingency tables. Pearson correlations were used for associations between continuous variables. Differences in SCFAs were evaluated using generalized linear models (GLMs), with covariate adjustment when appropriate (age, sex, BMI, metabolic syndrome, smoking or medications). Multiple testing was controlled using false discovery rate (FDR) correction.

Binary logistic regression was used to assess the ability of biomarkers to discriminate MDD from controls (controls as the reference group), adjusting for sex, BMI, and years of education. We reported Nagelkerke’s pseudo-R², Wald statistics with p-values, odds ratios (ORs) with 95% confidence intervals (CIs), and unstandardized coefficients (B) with standard errors (SE). To address class imbalance, we oversampled the control group by random replication. Model generalizability and stability were examined using 10-fold cross-validated stepwise linear discriminant analysis (LDA). Model performance was evaluated using classification accuracy, the maximum Kolmogorov–Smirnov statistic, the area under the ROC curve (AUC), the Gini index, and related diagnostic statistics.

Finally, manual multivariable regression analyses were conducted to identify predictors of API, EGF, and clinical phenotypes (OSOD, physiosomatic symptoms, current SI, and ROI). Candidate predictors included demographics, SCFAs, immune-inflammatory biomarkers, and ACEs. Forward stepwise regression was applied (entry p=0.05; removal p=0.07). We reported standardized β coefficients, degrees of freedom, p-values, R², and F statistics. Heteroscedasticity was assessed using White’s test and the modified Breusch–Pagan test. Multicollinearity was examined using tolerance and variance inflation factors (VIFs). All tests were two-tailed with α=0.05.

## Results

### Sociodemographic and clinical characteristics

**Table 1** summarizes sociodemographic and clinical characteristics. MDD and control groups did not differ in age, sex, years of education, smoking status, BMI, WC, MetS prevalence, or MetS ranking. Sexual abuse scores, Four ACEs, ROI, OSOD, physiosomatic symptoms, and current SI were higher in MDD than in controls.

**Table 1.** Socio-demographic, clinical, and immune-inflammation data of patients with major depression (MDD) and healthy controls (HC).

| Variables | HC (n = 38) | MDD (n = 102 ) | F/$\chi^2$ | df | P |
| --- | --- | --- | --- | --- | --- |
| Age (years) | 36.11 (13.41) | 35.78 (12.37) | 0.018 | 1/138 | 0.894 |
| Gender (Female/Male) | 25/13 | 71/31 | 0.187 | 1 | 0.665 |
| Education (years) | 14.26 (4.09) | 13.31 (3.43) | 1.903 | 1/138 | 0.170 |
| Smoking (yes/no) | 3/35 | 21/81 | FET | - | 0.084 |
| BMI (kg/m<sup>2</sup>) | 23.48 (4.12) | 22.30 (3.42) | 2.941 | 1/138 | 0.089 |
| Waist circumference (cm) | 79.76 (11.90) | 78.06 (11.69) | 0.579 | 1/138 | 0.448 |
| Mets (yes/no) | 10/28 | 20/81 | 0.692 | 1 | 0.405 |
| Ranking metabolic syndrome | 1.61 (1.48) | 1.47 (1.30) | 0.296 | 1/137 | 0.587 |
| Sex abuse (z score) | 0.38 (0.17) | 0.53 (0.27) | 11.510 | 1/138 | <0.001 |
| Four ACEs (z score) | 0.29 (0.25) | 0.56 (0.27) | 28.484 | 1/138 | <0.001 |
| ROI (z score) | -1.05 (0.15) | 0.27 (0.85) | MWUT | - | <0.001 |
| Current-SI (z score) | -0.77 (0.27) | 0.19 (0.98) | MWUT | - | <0.001 |
| Physiosomatic symptoms (z score) | -1.31 (0.43) | 0.42 (0.63) | MWUT | - | <0.001 |
| OSOD (z score) | -1.47 (0.27) | 0.46 (0.48) | MWUT | - | <0.001 |
| Acute phase-inflammatory index (z score) | -1.56 (1.97) | 0.48 (1.73) | MWUT | - | <0.001 |
| Th1 profile (z score) | -0.03 (0.92) | -0.01 (1.05) | MWUT | - | 0.722 |
| Th17 profile (z score) | -0.20 (0.94) | 0.04 (1.00) | MWUT | - | 0.193 |
| Th2 profile (z score) | -0.13 (1.00) | -0.05 (1.01) | MWUT | - | 0.514 |
| IRS profile (z score) | -0.07 (0.81) | -0.01 (1.05) | MWUT | - | 0.903 |
| CIRS profile (z score) | -0.32 (1.03) | 0.08 (0.99) | MWUT | - | 0.028 |
| EGF (z score) | -0.55 (0.87) | 0.17 (0.97) | MWUT | - | <0.001 |

Results are shown as mean (S.D.). Results of analysis of variance and analysis of contingency tables.
FET, Fisher's exact probability test; MWUT: Mann-Whitney U test; BMI, body mass index; Mets: metabolic syndrome. ACE, Adverse childhood experiences; SI, suicidal ideation; ROI, recurrence of illness; OSOD, overall severity of depression; Th, T helper; IRS, immune-inflammatory response system; CIRS, compensatory immunoregulatory response system; EGF, epidermal growth factor.

### Immune-inflammatory biomarkers and SCFAs in MDD vs controls

As shown in **Table 1**, the API, CIRS, and EGF were higher in MDD than in controls, whereas Th1, Th2, Th17, and IRS did not differ between groups. **Table 2** shows that acetic acid (AA), propionic acid (PA), butyric acid (BA), and protective SCFAs were lower in MDD, while isovaleric acid (isoVA), 2-methylbutyric acid (2-MBA), and the BSCFA index were higher. Isobutyric acid (isoBA), pentanoic acid, hexanoic acid, succinic acid, and neutral SCFAs did not differ between groups. These findings remained significant after FDR correction.

**Table 2.** Differences in short-chain fatty acids between major depressive disorder (MDD) and healthy controls (HC).

| Variables | HC (n = 38 ) | MDD (n = 102 ) | F value (df = 1/136) | P | q (FDR) |
| --- | --- | --- | --- | --- | --- |
| Acetic acid (µg/g) | 3894.00 (239.71) | 2887.14 (145.59) | 14.825 | <0.001 | **0.004** |
| Propionic acid (µg/g) | 2029.72 (160.58) | 1712.20 (97.53) | 5.894 | 0.017 | **0.029** |
| Butyric acid (µg/g) | 1940.50 (175.45) | 1559.51 (106.56) | 6.777 | 0.010 | **0.024** |
| Isobutyric acid (µg/g) | 226.28 (32.70) | 241.20 (19.86) | 0.382 | 0.538 | 0.587 |
| Isovaleric acid (µg/g) | 92.00 (29.84) | 170.17 (18.12) | 5.806 | 0.017 | **0.029** |
| Pentanoic acid (µg/g) | 244.14 (34.54) | 214.88 (20.98) | 1.342 | 0.249 | 0.332 |
| Hexanoic acid (µg/g) | 32.67 (8.62) | 21.75 (5.24) | 0.031 | 0.860 | 0.860 |
| 2-Methylbutyric acid (µg/g) | 79.53 (20.25) | 149.49 (12.30) | 7.982 | 0.005 | **0.015** |
| Succinic acid (µg/g) | 87.60 (32.81) | 63.77 (19.93) | 3.226 | 0.075 | 0.112 |
| Protective SCFAs (z score) | 1.19 (0.42) | -0.44 (0.26) | 10.737 | 0.001 | **0.004** |
| Neutral SCFAs (z score) | -0.61 (0.66) | 0.23 (0.40) | 1.179 | 0.279 | 0.335 |
| BSCFA index (z score) | -0.59 (0.15) | 0.22 (0.09) | 20.645 | <0.001 | **0.004** |

All data are shown as marginal estimated mean (SE). Results of GLM analysis with age and body mass index as covariates (sex and smoking are not significant). All significant differences shown in this table remained significant after False Discovery Rate p correction.
SCFA, short-chain fatty acids; BSCFA, branched-chain short-chain fatty acids.

### Effects of medications on SCFAs

Among all subjects, 56 received selective serotonin reuptake inhibitors (SSRIs), 15 serotonin-norepinephrine reuptake inhibitors (SNRIs), 16 5-HT1A agents, 52 benzodiazepines, 7 mood stabilizers, and 36 antipsychotics. The impacts of SNRI, 5-HT1A agents, and benzodiazepines on SCFA levels are summarized in **ESF Table 3**. After applying FDR correction, the use of 5-HT1A agents was modestly associated with higher levels of isoVA and the BSCFA index. Furthermore, after adjusting for the use of 5-HT1A agents, MDD diagnosis remained significantly associated with levels of AA, PA, BA, 2-MBA, protective SCFAs, and the BSCFA index. These findings suggest that the 5-HT1A agent-related effects and MDD-related effects on SCFAs were independent.

Notably, no SCFA variables remained significantly associated with SNRI and benzodiazepine use after FDR adjustment. SSRIs, mood stabilizers, and antipsychotic drugs showed no significant effects on any SCFA measures, even without FDR correction.

### Diagnostic performance of SCFAs and immune-inflammatory biomarkers

**Table 3** presents the binary logistic regression analyses evaluating the diagnostic accuracy of SCFAs and immune-inflammatory biomarkers for discriminating MDD patients from HCs. In the SCFA-based Model #1, MDD was significantly associated with lower AA and a higher BSCFA index (χ²=65.522, df=2, p<0.001; Nagelkerke R²=0.349). As shown in **ESF Table 4**, this model achieved acceptable overall classification accuracy.

**Table 3.** Results of binary logistic regression analyses with major depression (MDD) as the dependent variable and healthy controls as the reference group.

| Dichotomies |  | Explanatory Variables | B | SE | Wald<br>(df=1) | P | OR | 95% CI |
| --- | --- | --- | --- | --- | --- | --- | --- | --- |
| MDD vs. HC | Model #1 | BSCFA index | 0.837 | 0.177 | 22.340 | <0.001 | 2.309 | 1.632; 3.266 |
|  |  | AA | -0.961 | 0.205 | 22.014 | <0.001 | 0.383 | 0.256; 0.572 |
|  | Model #2 | BSCFA index | 0.639 | 0.200 | 10.216 | <0.001 | 1.894 | 1.280; 2.801 |
|  |  | AA | -0.952 | 0.259 | 13.495 | <0.001 | 0.386 | 0.232; 0.641 |
|  |  | API | 0.540 | 0.114 | 22.424 | <0.001 | 1.715 | 1.372; 2.145 |
|  |  | EGF | 0.785 | 0.245 | 10.258 | <0.001 | 2.193 | 1.356; 3.547 |
|  |  | Th2 | -0.595 | 0.221 | 7.265 | 0.007 | 0.552 | 0.358; 0.850 |
|  | Model #3 | BSCFA index | 0.519 | 0.216 | 5.754 | 0.016 | 1.680 | 1.100; 2.567 |
|  |  | AA | -1.083 | 0.294 | 13.538 | <0.001 | 0.339 | 0.190; 0.603 |
|  |  | API | 0.462 | 0.123 | 14.047 | <0.001 | 1.587 | 1.246; 2.020 |
|  |  | EGF | 1.078 | 0.285 | 14.267 | <0.001 | 2.940 | 1.680; 5.144 |
|  |  | Th2 | -0.748 | 0.256 | 8.539 | 0.003 | 0.473 | 0.287; 0.782 |
|  |  | Sex abuse | 0.701 | 0.276 | 6.472 | 0.011 | 2.016 | 1.175; 3.460 |
|  |  | Four ACEs | 0.644 | 0.236 | 7.432 | 0.006 | 1.905 | 1.199; 3.027 |

AA, acetic acid; API, acute phase inflammatory index; Th, T helper; EGF, epidermal growth factor; ACE, Adverse childhood experiences; BSCFA, branched-chain short-chain fatty acids.

After adding immune-inflammatory biomarkers (Model #2), predictive performance improved. Specifically, MDD was significantly associated with lower AA and Th2 levels and higher BSCFA index, API, and EGF. **ESF Table 4** further indicated increased classification accuracy for this expanded model. Stepwise LDA identified the same discriminant variables, which significantly separated MDD from HCs (Wilk’s λ=0.594, χ²=106.082, df=5, p<0.001; canonical correlation=0.637): BSCFA, AA, API, EGF, and Th2. **Figure 1** shows the receiver operating characteristic (ROC) curve for this model, yielding an AUC of 0.874 (±0.024) and a Gini index of 0.749, with a 10-fold cross-validated sensitivity of 76.3% and specificity of 81.1%.

**Figure 1.**
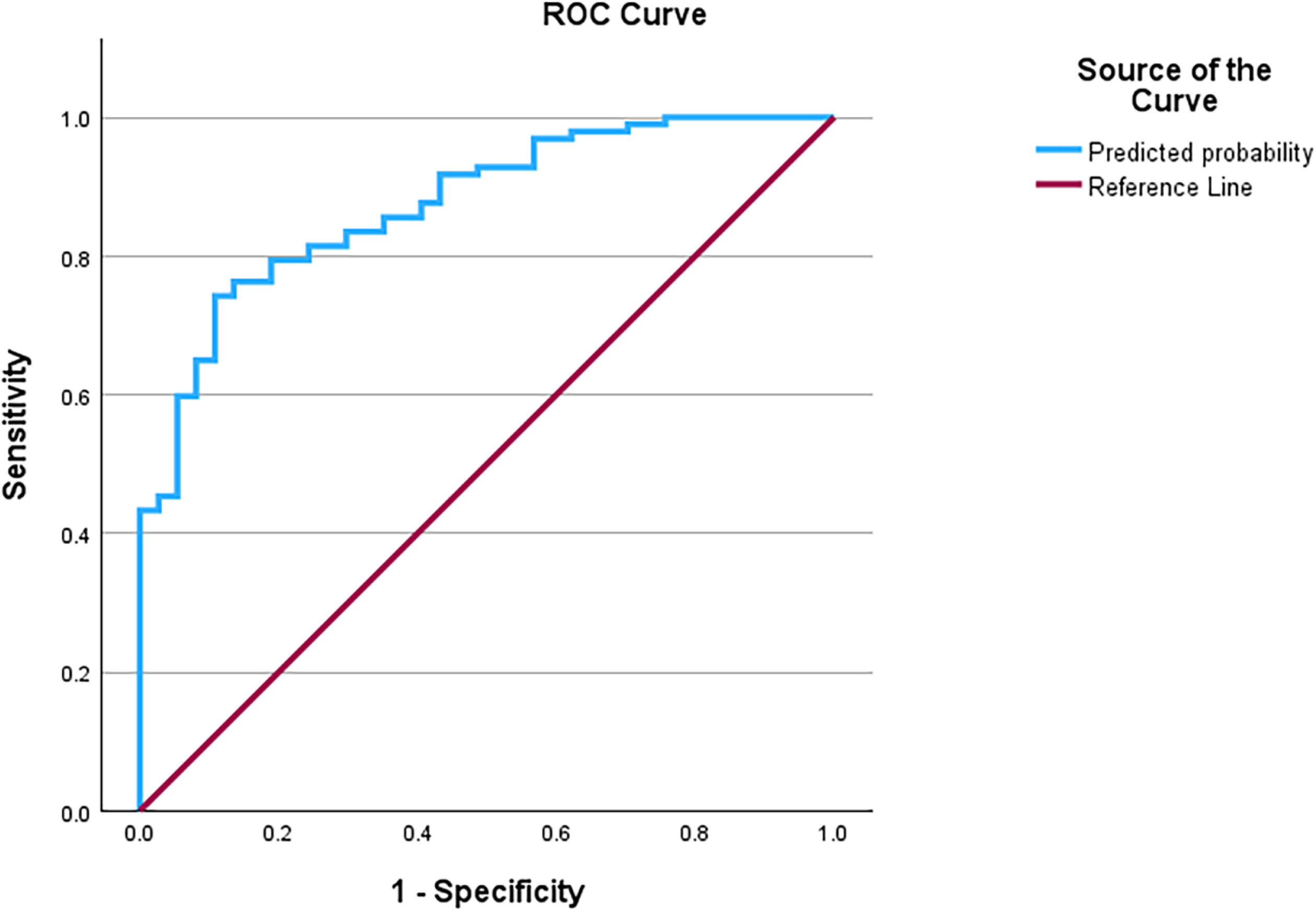
The receiving operating curve discriminating major depressive disorder from controls using BSCFA index, acetic acid, acute phase inflammatory index, epidermal growth factor and T helper 2 as explanatory variables. BSCFA, branched-chain short-chain fatty acids.

In Model #3, ACEs were additionally incorporated. The final model included the four ACEs and sexual abuse scores, together with the same biomarkers as in Model #2. As shown in **ESF Table 4**, this further improved predictive ability and overall classification accuracy.

### Correlations between SCFAs, ACEs, clinical phenotypes, and immune-inflammatory biomarkers

**Table 4** reports Pearson correlations between representative SCFAs and ACEs, immune-inflammatory biomarkers, and clinical phenotypes. The four ACE score was negatively correlated with AA and PA, whereas sexual abuse was positively correlated with the BSCFA index. OSOD, physiosomatic symptoms, and ROI were significantly negatively correlated with AA, PA, and BA and positively correlated with the BSCFA index. The current SI was negatively correlated with AA.

**Table 4.** Intercorrelation matrix (Pearson’s correlation coefficients) between major short-chain fatty acids, adverse childhood experiences (ACEs), immune-inflammation biomarkers and the clinical phenome of depression.

| Variables | AA | PA | BA | BSCFA index | SA |
| --- | --- | --- | --- | --- | --- |
| Sex abuse | -0.003 | -0.054 | -0.093 | 0.200* | -0.134 |
| Four ACEs | -0.209* | -0.167* | -0.154 | 0.077 | -0.053 |
| API | -0.270** | -0.118 | -0.212* | 0.254** | -0.184* |
| EGF | -0.253** | -0.141 | -0.139 | 0.208* | -0.162 |
| Th1 | -0.036 | 0.001 | -0.028 | 0.025 | -0.092 |
| Th17 | -0.045 | -0.015 | 0.012 | 0.038 | -0.146 |
| Th2 | -0.088 | -0.11 | -0.01 | 0.1 | -0.195* |
| IRS | -0.029 | 0.02 | -0.044 | 0.053 | -0.12 |
| CIRS | -0.151 | -0.111 | -0.056 | 0.182* | -0.171* |
| OSOD | -0.368*** | -0.253** | -0.249** | 0.364*** | -0.158 |
| Physiosomatic symptoms | -0.293*** | -0.213* | -0.197* | 0.314*** | -0.114 |
| Current SI | -0.209* | -0.108 | -0.091 | 0.168 | -0.077 |
| ROI | -0.343*** | -0.211* | -0.179* | 0.205* | -0.13 |

\* Correlation is significant at the 0.05 level (2-tailed). \*\* Correlation is significant at the 0.01 level (2-tailed). \*\*\* Correlation at 0.001(2-tailed).
AA, acetic acid; PA, propionic acid; BA, butyric acid; SA, succinic acid; ACE, Adverse childhood experiences; API, acute phase inflammatory index; Th, T helper; EGF, epidermal growth factor; IRS, immune-inflammatory response system; CIRS, compensatory immunoregulatory response system; OSOD, overall severity of depression; SI, suicidal ideation; ROI, recurrence of illness; BSCFA, branched-chain short-chain fatty acids.

API was significantly negatively correlated with AA, PA, BA, and succinic acid, and positively correlated with the BSCFA index. EGF was significantly negatively correlated with AA and positively correlated with the BSCFA index. In addition, Th2 and CIRS were negatively correlated with succinic acid, and CIRS was positively correlated with the BSCFA index.

### SCFA predictors of API and EGF

**Table 5** (Model#1 - Model#2) shows the multivariable regression analyses with API and EGF as dependent variables and SCFAs as predictors, controlling for sex, age, and BMI. In Model #1, 12.5% of the variance in API was explained by the BSCFA index (positive) and AA (negative). In Model #2, 9.7% of the variance in EGF was explained by the same two predictors.

**Table 5.** Results of multiple regression analysis with severity of depression score, API and EGF as dependent variables.

| Dependent variables | Explanatory variables | Parameter estimates + statistics |  |  | Model statistics + effect sizes |  |  |  |
| --- | --- | --- | --- | --- | --- | --- | --- | --- |
| | | $\beta$ | T | p | F<sub>model</sub> | df | p | R<sup>2</sup> |
| #1. API | **Model** |  |  |  | **9.714** | **2/136** | **<0.001** | **0.125** |
|  | AA | -0.247 | -3.063 | 0.003 |  |  |  |  |
|  | BSCFA index | 0.230 | 2.850 | 0.005 |  |  |  |  |
| #2. EGF | **Model** |  |  |  | **6.997** | **2/131** | **0.001** | **0.097** |
|  | AA | -0.233 | -2.782 | 0.006 |  |  |  |  |
|  | BSCFA index | 0.182 | 2.173 | 0.032 |  |  |  |  |
| #3. OSOD (no ACEs and immune-inflammation biomarkers entered) | **Model** |  |  |  | **19.801** | **2/125** | **<0.001** | **0.241** |
|  | AA | -0.331 | -4.225 | <0.001 |  |  |  |  |
|  | BSCFA index | 0.326 | 4.157 | <0.001 |  |  |  |  |
| #4. OSOD (after ACEs and immune-inflammation biomarkers entered) | **Model** |  |  |  | **24.487** | **4/118** | **<0.001** | **0.454** |
|  | API | 0.315 | 4.253 | <0.001 |  |  |  |  |
|  | Four ACEs | 0.32 | 4.443 | <0.001 |  |  |  |  |
|  | BSCFA index | 0.293 | 4.126 | <0.001 |  |  |  |  |
|  | Propionic acid | -0.173 | -2.429 | 0.017 |  |  |  |  |
| #5. Physiosomatic symptoms (no ACEs and immune-inflammation biomarkers entered) | **Model** |  |  |  | **12.466** | **2/125** | **<0.001** | **0.166** |
|  | BSCFA index | 0.351 | 4.257 | <0.001 |  |  |  |  |
|  | Propionic acid | -0.263 | -3.188 | 0.002 |  |  |  |  |
| #6. Physiosomatic symptoms (after ACEs and immune-inflammation biomarkers entered) | **Model** |  |  |  | **15.645** | **5/117** | **<0.001** | **0.401** |
|  | API | 0.289 | 3.744 | <0.001 |  |  |  |  |
|  | Four ACEs | 0.252 | 3.124 | 0.002 |  |  |  |  |
|  | BSCFA index | 0.261 | 3.284 | 0.001 |  |  |  |  |
|  | Pentanoic acid | -0.186 | -2.394 | 0.018 |  |  |  |  |
|  | Sex abuse | 0.17 | 2.171 | 0.032 |  |  |  |  |
| #7. Current SI (no ACEs and immune-inflammation biomarkers entered) | **Model** |  |  |  | **5.867** | **1/129** | **0.017** | **0.044** |
|  | AA | -0.209 | -2.422 | 0.017 |  |  |  |  |
| #8. ROI (no ACEs and immune-inflammation biomarkers entered) | **Model** |  |  |  | **10.885** | **2/128** | **<0.001** | **0.145** |
|  | AA | -0.324 | -3.935 | <0.001 |  |  |  |  |
|  | BSCFA index | 0.168 | 2.037 | 0.044 |  |  |  |  |
| #9. ROI (after ACEs and immune-inflammation biomarkers entered) | **Model** |  |  |  | **18.647** | **4/121** | **<0.001** | **0.381** |
|  | Four ACEs | 0.421 | 5.513 | <0.001 |  |  |  |  |
|  | API | 0.187 | 2.377 | 0.019 |  |  |  |  |
|  | AA | -0.195 | -2.561 | 0.012 |  |  |  |  |
|  | Th17 | 0.171 | 2.354 | 0.020 |  |  |  |  |

API, acute phase inflammatory index; AA, acetic acid; EGF, epidermal growth factor; OSOD, overall severity of depression; ACE, Adverse childhood experiences; SI, suicidal ideation; ROI, recurrence of illness; Th, T helper; IRS, immune-inflammatory response system; MDD, major depressive disorder; BSCFA, branched-chain short-chain fatty acids.

### Predictors of clinical phenotypes

**Table 5** summarizes multivariable regression models predicting MDD clinical phenotypes. In Model #3, 24.1% of the variance in OSOD was explained by the BSCFA index (positive) and AA (negative). After accounting for ACEs and immune-inflammatory biomarkers (Model #4), 45.4% of OSOD variance was explained by API, Four ACEs, and the BSCFA index (all positive) and by PA (negative). For physiosomatic symptoms, the SCFA-only Model #5 explained 16.6% of variance, with the BSCFA index positively and PA negatively associated. After including ACEs and immune-inflammatory biomarkers (Model #6), 40.1% of variance was explained by API, the BSCFA index, Four ACEs, and sexual abuse (all positive), and by pentanoic acid (negative). For current SI, the SCFA-only Model #7 explained 4.4% of variance, with AA showing a negative contribution. After incorporating ACEs and immune-inflammatory biomarkers, the SCFA effects were no longer significant.

For ROI, the SCFA-only Model #8 explained 14.5% of variance, with the BSCFA index positively and AA negatively associated. After including ACEs and immune-inflammatory biomarkers (Model #9), 38.1% of ROI variance was explained by Four ACEs, API, and Th17 (all positive), and by AA (negative). In analyses restricted to the MDD cohort, no SCFA effects could be observed.

## Discussion

This study provides comprehensive evidence for the interplay between gut metabolic dysregulation and systemic immune activation in MDD, further reinforcing the critical role of the NIMETOX pathway in its pathophysiology. Our core findings include: (1) MDD patients exhibit a distinct metabolic profile characterized by a significant depletion of protective straight-chain saccharolytic SCFAs (AA, PA, BA) and a marked elevation in BSCFAs (e.g., isoVA, 2-MBA); (2) this metabolic imbalance is closely associated with systemic immune activation, indicated by elevated API and EGF; (3) a multidimensional model combining low AA, high BSCFAs, and immune markers demonstrates adequate diagnostic accuracy for MDD (AUC = 0.874); (4) alterations in the SCFA profile—particularly decreased AA/PA and an elevated BSCFA index—are robust predictors of clinical phenotypes, including OSOD, physiosomatic symptoms, and ROI; and (5) the use of specific antidepressive medications (notably 5-HT1A agonists) may alter the SCFA profile independently of the disease, potentially exacerbating the elevation of BSCFAs.

### SCFAs in MDD: A Pathological Shift from Saccharolytic to Proteolytic Fermentation

The observed depletion of protective straight-chain SCFAs, particularly BA, aligns closely with previous reports of reduced butyrate-producing bacteria (e.g., *Faecalibacterium* and *Coprococcus*) in MDD (4, 14, 15). This depletion essentially reflects a decline in the saccharolytic fermentation capacity of the gut microbiota. More breakthrough evidence lies in the significant elevation of BSCFAs, which provides direct metabolic proof of a pathological shift toward "proteolytic fermentation" within the gut of MDD patients. This transition is likely driven by the expansion of proteolytic taxa frequently enriched in MDD, such as *Bacteroides*, *Alistipes*, and *Clostridium* (4, 14, 15, 21). This metabolic reprogramming constitutes a "double hit" to the host: first, the loss of beneficial straight-chain SCFAs deprives the intestinal barrier of essential energy and repair mechanisms; second, the co-production of cytotoxic proteolytic metabolites (e.g., p-cresol, indoles, and ammonia) alongside BSCFAs directly damages the intestinal epithelium, thereby driving systemic inflammation and oxidative stress (18-20, 22-24). Consequently, this metabolic shift likely acts as a primary upstream driver of the oxidative and nitrosative stress described in the NIMETOX pathway (3, 20).

### SCFAs and Systemic Immune Activation (API/EGF)

Our data reveals a tight mechanistic link between the depletion of protective SCFAs, the accumulation of BSCFAs, and systemic immune activation (elevated API and EGF). This relationship can be delineated across multiple dimensions, including gut barrier integrity, immune tolerance, and vagal tone.

First, the intestinal mucosa suffers from "energy starvation" and toxic disruption. As previously noted, AA and BA are indispensable for maintaining epithelial tight junctions (16, 17). Their substantial depletion deprives the physical barrier of crucial energetic and signaling support (18, 19). Concurrently, proteolytic metabolites acting as "metabolic disruptors" alter the luminal pH and interfere with colonocyte respiration (22-24). This synergistic effect of lost protection and active disruption likely impairs tight junctions, thereby increasing intestinal permeability and facilitating the pathological translocation of endotoxins (e.g., LPS) into the systemic circulation (5). Translocated LPS subsequently triggers targeted humoral immune responses and initiates the "TLR-radical cycle" (5-7, 20). The resulting surge in inflammatory mediators can reflexively exacerbate barrier damage via the JNK signaling pathway, thus forming a vicious cycle (50, 51).

Second, the loss of immune tolerance converges with the failure of the cholinergic anti-inflammatory pathway (CAP). The deficit in BA—a natural HDAC inhibitor—attenuates HDAC inhibition within immune cells, impairing the differentiation of regulatory T cells (Tregs) and leaving the immune system in a hyper-reactive state (52, 53). Simultaneously, the diminished vagal tone frequently observed in MDD suggests an impairment of the CAP, which normally suppresses intestinal inflammation via efferent vagal fibers (54-56). Under these conditions, gut-derived pro-inflammatory processes propagate unchecked.

Furthermore, while succinate is recognized as a key immunometabolic signal capable of stabilizing HIF-1α and promoting inflammation under specific conditions (32), our results showed no significant group differences and a negative correlation with the API. This strongly implies that in the specific pathological context of MDD, systemic inflammation is predominantly driven by the combined deficit of protective SCFAs and the excess of toxic BSCFAs, rather than by luminal succinate accumulation alone.

### Diagnostic Utility and Clinical Phenotypes: Defining a "Gut-Immune Biotype"

A pivotal outcome of this study is the high diagnostic accuracy achieved by combining SCFA profiles with immune markers. Our multidimensional model (incorporating low AA, high BSCFAs, high API/EGF, and low Th2) exhibited adequate discriminative power (AUC = 0.874), indicating that MDD is a syndrome characterized by the co-occurrence of metabolic and immune perturbations. This concurrent profile delineates a distinct "gut-immune biotype." In clinical practice, identifying patients with this biotype could advance precision medicine by prioritizing them for immune therapies or microbiome-targeted interventions (e.g., precise pre/probiotics) over standard monoaminergic treatments.

Beyond diagnostic value, alterations in the SCFA profile strongly predicted specific clinical phenotypes. Modeling revealed that an elevated BSCFA index and decreased protective SCFAs (AA/PA) are significant predictors of higher OSOD, more severe physiosomatic symptoms, and an increased ROI. These macroscopic clinical associations are grounded in profound microscopic neurobiological mechanisms:

*Energy Deficits Drive Fatigue and Psychomotor Retardation:* AA exhibits the highest blood-brain barrier (BBB) penetrance among SCFAs and is a critical substrate for astrocytes and microglia (such as in oxidative phosphorylation) (57, 58). Its depletion in the gut equates to a loss of vital energetic support for the CNS, potentially explaining the profound fatigue and psychomotor retardation typical of MDD.

*Neurochemical Hijacking and Central Over-Inhibition:* Systemic inflammation caused by intestinal metabolic disorders potently activates IDO, initiating the "tryptophan steal" or kynurenine pathway. This not only deprives the body of the precursor for serotonin synthesis, but also leads to excessive production of neurotoxic metabolites (such as quinolinic acid), directly exacerbating the core symptoms of depression (3, 4, 59, 60). Moreover, the significantly elevated BSCFAs (particularly isoVA) share structural similarities with the inhibitory neurotransmitter GABA and the antiepileptic drug valproate (61). We hypothesize that excess BSCFAs may cross a compromised BBB and act as "central depressants," interfering with normal neuronal excitability. This offers a novel neurochemical explanation for symptoms like hypersomnia and cognitive blunting.

*Aberrant Visceral Afferent Signaling Exacerbates Somatic Distress:* Unlike straight-chain SCFAs that transmit neuroprotective signals, high concentrations of BSCFAs and associated putrefactive products acting as noxious stimuli. They may hyper-activate vagal afferent chemoreceptors, continuously relaying signals of visceral stress and metabolic toxicity to the brainstem (4, 62). This aberrant bottom-up signaling likely contributes to central sensitization and visceral hypersensitivity, exacerbating somatic pain and gastrointestinal discomfort.

*SCFA Dysregulation Predicts ROI:* Chronic exposure to a "low protective/high toxic" SCFA state not only compromises energy supply to colonocytes but also disrupts the signaling functions of SCFAs in modulating immune homeostasis and hippocampal neuroplasticity through activation of FFAR2/3 and inhibition of HDACs (63-66). The loss of these protective mechanisms significantly reduces individual resilience to stressful events, potentially contributing to recurrent episodes of depression. Indeed, in chronic unpredictable mild stress animal models, exogenous sodium butyrate supplementation has been demonstrated to effectively reverse depressive-like behaviors by inhibiting HDACs, upregulating brain-derived neurotrophic factor expression, and repairing blood-brain barrier integrity (67, 68).

Collectively, these multidimensional neurobiological perturbations are not isolated events but represent the direct clinical manifestations of the intertwining of "gut metabolic toxicity" and "systemic neuroinflammation" within the NIMETOX cycle.

### Potential Effects of Specific Antidepressants (5-HT1A Agonists) on SCFAs

Our in-depth analysis of medication effects revealed that among various drugs, only 5-HT1A agonists remained significantly associated with elevated isoVA and BSCFA index after FDR correction. This suggests that 5-HT1A agents may act as "ecological stressors" with non-antibiotic antimicrobial properties within the gut (69). These medications might alter gut motility or inhibit specific butyrate-producing taxa, inadvertently selecting for resistant strains that mediate proteolytic fermentation (70, 71). This drug-induced dysbiosis implies that while certain medications may alleviate monoamine-related symptoms, they may simultaneously perpetuate—or even exacerbate—underlying metabolic dysregulation at the microecological level. Future treatment strategies must therefore carefully evaluate the off-target effects of antidepressants on the microbiome.

## Limitations

It must be acknowledged that this study has some limitations. The cross-sectional design precludes causal inferences. Fecal SCFAs serve only as a proxy for gut metabolism and cannot precisely reflect systemic absorption rates. Additionally, the lack of detailed dietary data and the relatively small sample size of the 5-HT1A subgroup necessitate further validation in large-scale, longitudinal cohorts. While our current cohort provided robust insights, these findings need to be cross-validated in independent cohorts from diverse ethnic and cultural backgrounds to establish their generalizability. Moving forward, future research should specifically focus on exploring the complex associations between SCFAs, peripheral NIMETOX biomarkers, and brain imaging data to further elucidate the underlying mechanisms.

## Conclusion

In summary, leveraging the NIMETOX theoretical framework, this study comprehensively elucidates a distinct "gut-immune biotype" in MDD. Our key findings demonstrate that MDD involves not only the depletion of protective saccharolytic metabolites (straight-chain SCFAs) but also a pathological shift toward "proteolytic fermentation" (marked elevation of toxic BSCFAs). This complex metabolic imbalance is intricately linked to systemic immune inflammation and serves as a robust predictor of OSOD, physiosomatic symptoms, and ROI. Furthermore, a multidimensional model integrating low AA, high BSCFAs, and immune markers (API/EGF) achieves adequate diagnostic efficacy. Notably, specific antidepressants (particularly 5-HT1A agonists) may act as additional ecological stressors, exacerbating the elevation of BSCFAs independently of the underlying disease. These findings underscore the urgent need to transition toward precision psychiatry. Capitalizing on this "proteolysis-inflammation" biosignature provides a compelling, objective rationale for identifying specific patient subpopulations and developing personalized therapeutic strategies aimed at rectifying aberrant gut fermentation patterns.

## Data Availability

The database created during this investigation will be provided by the corresponding author (MM) upon a reasonable request once the authors have thoroughly used the data set.

## Ethics approval

This study was approved by the ethics committee of Sichuan Provincial People’s Hospital [Ethics (Research) 2024-203] and strictly followed ethical and privacy regulations.

## Consent to participate

Before participating in this study, each subject provided written informed consent.

## Consent for publication

All authors have given their approval for this paper to be published.

## Author’s contributions

Mengqi Niu: visualization, writing - original draft, statistical analyses, recruiting participants. Yiping Luo, Chenkai Yangyang, Jing Li: recruiting participants. Abbas F. Almulla, Andre F. Carvalho: writing - review and editing. Yingqian Zhang: conceptualization, writing - review and editing. Michael Maes: supervision, conceptualization, formal analysis, writing - review and editing. All authors approved the submitted manuscript.

## ELECTRONIC SUPPLEMENTARY FILE (ESF)

**ESF, Table 1.**
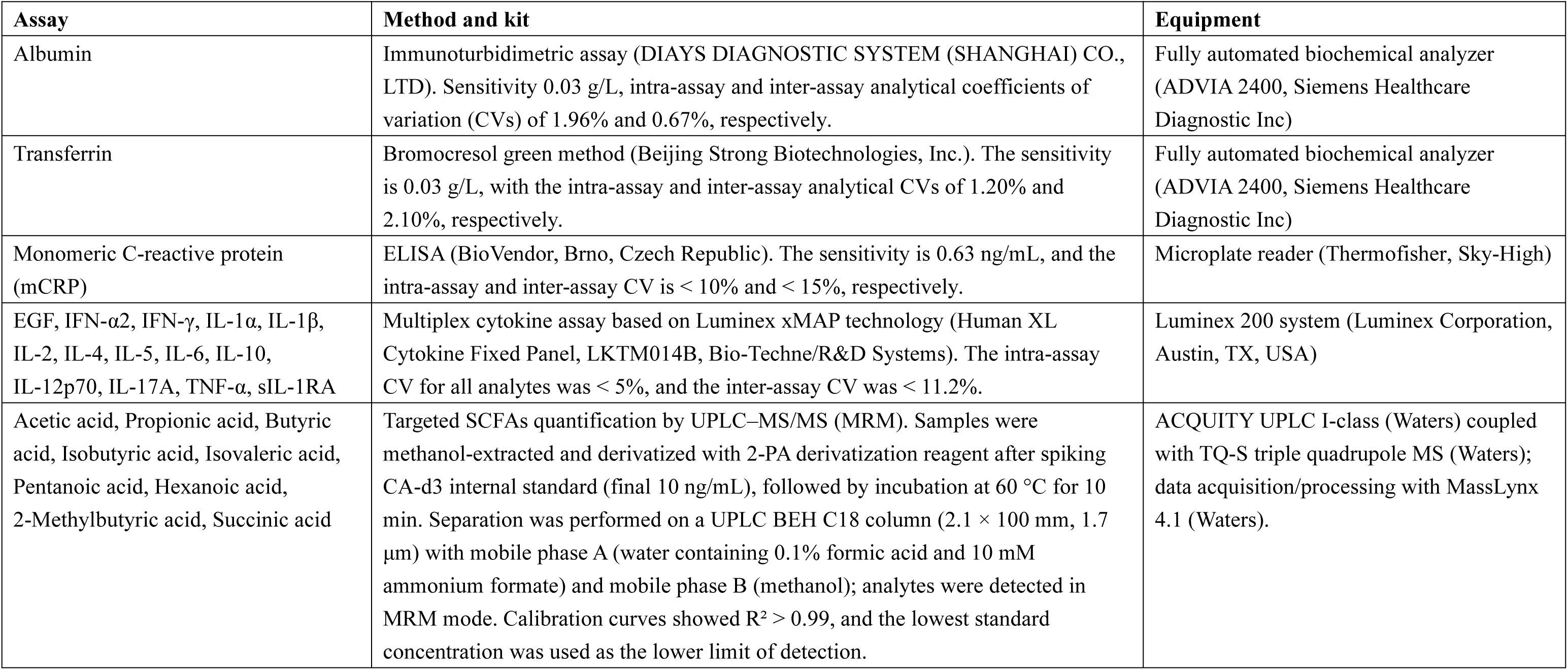
Methods used to assay the biomarkers in the present study.

| Assay | Method and kit | Equipment |
| --- | --- | --- |
| Albumin | Immunoturbidimetric assay (DIAYS DIAGNOSTIC SYSTEM (SHANGHAI) CO., LTD). Sensitivity 0.03 g/L, intra-assay and inter-assay analytical coefficients of variation (CVs) of 1.96% and 0.67%, respectively. | Fully automated biochemical analyzer (ADVIA 2400, Siemens Healthcare Diagnostic Inc) |
| Transferrin | Bromocresol green method (Beijing Strong Biotechnologies, Inc.). The sensitivity is 0.03 g/L, with the intra-assay and inter-assay analytical CVs of 1.20% and 2.10%, respectively. | Fully automated biochemical analyzer (ADVIA 2400, Siemens Healthcare Diagnostic Inc) |
| Monomeric C-reactive protein (mCRP) | ELISA (BioVendor, Brno, Czech Republic). The sensitivity is 0.63 ng/mL, and the intra-assay and inter-assay CV is < 10% and < 15%, respectively. | Microplate reader (Thermofisher, Sky-High) |
| EGF, IFN-α2, IFN-γ, IL-1α, IL-1β, IL-2, IL-4, IL-5, IL-6, IL-10, IL-12p70, IL-17A, TNF-α, sIL-1RA | Multiplex cytokine assay based on Luminex xMAP technology (Human XL Cytokine Fixed Panel, LKTM014B, Bio-Techne/R&D Systems). The intra-assay CV for all analytes was < 5%, and the inter-assay CV was < 11.2%. | Luminex 200 system (Luminex Corporation, Austin, TX, USA) |
| Acetic acid, Propionic acid, Butyric acid, Isobutyric acid, Isovaleric acid, Pentanoic acid, Hexanoic acid, 2-Methylbutyric acid, Succinic acid | Targeted SCFAs quantification by UPLC–MS/MS (MRM). Samples were methanol-extracted and derivatized with 2-PA derivatization reagent after spiking CA-d3 internal standard (final 10 ng/mL), followed by incubation at 60 °C for 10 min. Separation was performed on a UPLC BEH C18 column (2.1 × 100 mm, 1.7 μm) with mobile phase A (water containing 0.1% formic acid and 10 mM ammonium formate) and mobile phase B (methanol); analytes were detected in MRM mode. Calibration curves showed $R^2 > 0.99$, and the lowest standard concentration was used as the lower limit of detection. | ACQUITY UPLC I-class (Waters) coupled with TQ-S triple quadrupole MS (Waters); data acquisition/processing with MassLynx 4.1 (Waters). |

**ESF, Table 2.** Description of the profiles used in this study.

**ESF, Table 2.** Description of the profiles used in this study.

| Profile | Members |
| --- | --- |
| **Acute phase-inflammatory index** | z(mCRP) - z(albumin) - z(transferrin) |
| **T helper (Th)-1** | IFN-α2, IFN-γ, IL-2, IL-12p70 |
| **Th-17** | IL-6, IL-17A |
| **Th-2** | IL-4, IL-5 |
| **IRS** | IL-1α, IL-1β, TNF-α, IL-6, Th1, and Th17 profiles |
| **CIRS** | IL-4, IL-5, IL-10, sIL-1RA |
| **Protective SCFAs** | z(acetic acid) + z(propionic acid) + z(butyric acid) |
| **Neutral SCFAs** | z(valeric acid) + z(isobutyric acid) + z(isovaleric acid) + z(hexanoic acid) + z(2-methylbutyric acid) |
| **BSCFA index** | z(isovaleric acid) + z(2-methylbutyric acid) - z(isobutyric acid) |

IRS: immune-inflammatory response system; CIRS: compensatory immunoregulatory system; BSCFA, branched-chain short-chain fatty acids.
Adapted from:
*Almulla AF, Abbas Abo Algon A, Tunvirachaisakul C, Al-Hakeim HK, Maes M. T helper-1 activation via interleukin-16 is a key phenomenon in the acute phase of severe, first-episode major depressive disorder and suicidal behaviors. J Adv Res. 2024 Oct;64:171-181. doi: 10.1016/j.jare.2023.11.012. Epub 2023 Nov 13. PMID: 37967811; PMCID: PMC11464466.*
*Maes M, Rachayon M, Jirakran K, Sodsai P, Klinchanhom S, Galecki P, Sughondhabiroom A, Basta-Kaim A. The Immune Profile of Major Dismood Disorder: Proof of Concept and Mechanism Using the Precision Nomothetic Psychiatry Approach. Cells. 2022 Mar 31;11(7):1183. doi: 10.3390/cells11071183. PMID: 35406747; PMCID: PMC8997660.*

**ESF, Table 3.** Effects of the use of SNRI, 5-HT1A agents, and benzodiazepines on the short-chain fatty acids in all cohorts.

| Dependent Variable | Effects of the use of drugs |  |  | Effects of MDD |  |  |
| --- | --- | --- | --- | --- | --- | --- |
|  | F | P | q (FDR) | F | P | q (FDR) |
| **SNRI** |  |  |  |  |  |  |
| Acetic acid (z score) | 1.059 | 0.305 | 0.523 | 17.348 | <0.001 | **0.004** |
| Propionic acid (z score) | 2.546 | 0.113 | 0.332 | 8.539 | 0.004 | **0.010** |
| Butyric acid (z score) | 1.936 | 0.166 | 0.332 | 9.658 | 0.002 | **0.006** |
| Isobutyric acid (z score) | 0.686 | 0.409 | 0.545 | 0.459 | 0.499 | 0.544 |
| Isovaleric acid (z score) | 0.201 | 0.655 | 0.715 | 5.388 | 0.022 | **0.038** |
| Pentanoic acid (z score) | 5.483 | 0.021 | 0.126 | 2.419 | 0.122 | 0.163 |
| Hexanoic acid (z score) | 7.689 | 0.006 | 0.072 | 0.147 | 0.702 | 0.702 |
| 2-Methylbutyric acid (z score) | 0.846 | 0.359 | 0.538 | 7.533 | 0.007 | **0.014** |
| Succinic acid (z score) | 0.090 | 0.765 | 0.765 | 3.472 | 0.065 | 0.098 |
| Protective SCFAs (z score) | 2.213 | 0.139 | 0.332 | 14.151 | <0.001 | **0.004** |
| Neutral SCFAs (z score) | 2.952 | 0.088 | 0.332 | 0.794 | 0.374 | 0.449 |
| BSCFA index (z score) | 0.267 | 0.606 | 0.715 | 19.057 | <0.001 | **0.004** |
| **5-HT1A** |  |  |  |  |  |  |
| Acetic acid (z score) | 0.612 | 0.435 | 0.729 | 14.157 | <0.001 | **0.004** |
| Propionic acid (z score) | 0.022 | 0.883 | 0.905 | 6.703 | 0.011 | **0.026** |
| Butyric acid (z score) | 0.169 | 0.682 | 0.845 | 7.293 | 0.008 | **0.024** |
| Isobutyric acid (z score) | 5.545 | 0.020 | 0.053 | 0.122 | 0.727 | 0.793 |
| Isovaleric acid (z score) | 7.330 | 0.008 | **0.048** | 3.534 | 0.062 | 0.093 |
| Pentanoic acid (z score) | 0.489 | 0.486 | 0.729 | 1.423 | 0.235 | 0.313 |
| Hexanoic acid (z score) | 2.744 | 0.100 | 0.200 | 0.027 | 0.869 | 0.869 |
| 2-Methylbutyric acid (z score) | 6.171 | 0.014 | 0.053 | 5.980 | 0.016 | **0.032** |
| Succinic acid (z score) | 0.014 | 0.905 | 0.905 | 3.799 | 0.053 | 0.091 |
| Protective SCFAs (z score) | 0.145 | 0.704 | 0.845 | 11.103 | 0.001 | **0.004** |
| Neutral SCFAs (z score) | 5.407 | 0.022 | 0.053 | 0.568 | 0.452 | 0.542 |
| BSCFA index (z score) | 7.723 | 0.006 | **0.048** | 15.818 | <0.001 | **0.004** |
| **Benzodiazepines** |  |  |  |  |  |  |
| Acetic acid (z score) | 2.074 | 0.152 | 0.182 | 18.108 | <0.001 | **0.003** |
| Propionic acid (z score) | 4.449 | 0.037 | 0.091 | 11.161 | 0.001 | **0.003** |
| Butyric acid (z score) | 1.264 | 0.263 | 0.287 | 9.385 | 0.003 | **0.007** |
| Isobutyric acid (z score) | 5.401 | 0.022 | 0.091 | 0.094 | 0.759 | 0.828 |
| Isovaleric acid (z score) | 3.135 | 0.079 | 0.129 | 1.891 | 0.171 | 0.228 |
| Pentanoic acid (z score) | 5.307 | 0.023 | 0.091 | 4.155 | 0.043 | 0.086 |
| Hexanoic acid (z score) | 3.256 | 0.073 | 0.129 | 0.444 | 0.506 | 0.607 |
| 2-Methylbutyric acid (z score) | 4.389 | 0.038 | 0.091 | 2.912 | 0.090 | 0.154 |
| Succinic acid (z score) | 0.071 | 0.791 | 0.791 | 2.612 | 0.108 | 0.162 |
| Protective SCFAs (z score) | 2.985 | 0.086 | 0.129 | 15.476 | <0.001 | **0.003** |
| Neutral SCFAs (z score) | 5.945 | 0.016 | 0.091 | 0.000 | 0.999 | 0.999 |
| BSCFA index (z score) | 2.215 | 0.139 | 0.182 | 11.379 | <0.001 | **0.003** |

All results of GLM analysis with MDD and the medications use as fixed factors. The bold font indicates that there is still a significant difference after FDR correction. SCFA, short-chain fatty acids; BSCFA, branched-chain short-chain fatty acids; SNRIs, serotonin-norepinephrine reuptake inhibitors.

**ESF Table 4.**
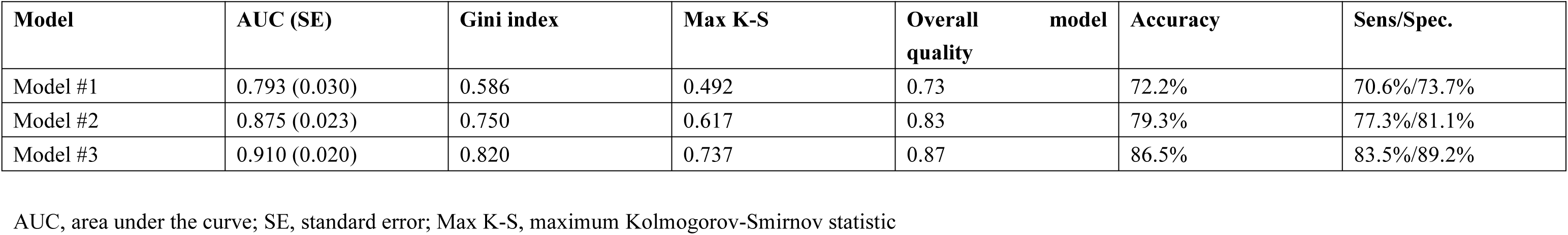
Performance evaluation metrics for different models (see Table 3).

